# TRACKING AND PREDICTING COVID-19 RADIOLOGICAL TRAJECTORY USING DEEP LEARNING ON CHEST X-RAYS: INITIAL ACCURACY TESTING

**DOI:** 10.1101/2020.05.01.20086207

**Authors:** S. Duchesne, D. Gourdeau, P. Archambault, C. Chartrand-Lefebvre, L. Dieumegarde, R. Forghani, C. Gagné, A. Hains, D. Hornstein, H. Le, S. Lemieux, M.H. Lévesque, D. Martin, L. Rosenbloom, A. Tang, F. Vecchio, O. Potvin, N. Duchesne

**Author notes:** Co-First Authors (S. Duchesne, D. Gourdeau). Corresponding author: Simon Duchesne, P.Eng., Ph.D., CERVO Brain Research Center, 2601 de la Canardière, Québec, Québec, Canada G1J 2G3.

## Abstract

**Background:** Decision scores and ethically mindful algorithms are being established to adjudicate mechanical ventilation in the context of potential resources shortage due to the current onslaught of COVID-19 cases. There is a need for a reproducible and objective method to provide quantitative information for those scores.

**Purpose:** Towards this goal, we present a retrospective study testing the ability of a deep learning algorithm at extracting features from chest x-rays (CXR) to track and predict radiological evolution.

**Materials and Methods:** We trained a repurposed deep learning algorithm on the CheXnet open dataset (224,316 chest X-ray images of 65,240 unique patients) to extract features that mapped to radiological labels. We collected CXRs of COVID-19-positive patients from two open-source datasets (last accessed on April 9, 2020)(Italian Society for Medical and Interventional Radiology and MILA). Data collected form 60 pairs of sequential CXRs from 40 COVID patients (mean age ± standard deviation: 56 ± 13 years; 23 men, 10 women, seven not reported) and were categorized in three categories: “Worse”, “Stable”, or “Improved” on the basis of radiological evolution ascertained from images and reports. Receiver operating characteristic analyses, Mann-Whitney tests were performed.

**Results:** On patients from the CheXnet dataset, the area under ROC curves ranged from 0.71 to 0.93 for seven imaging features and one diagnosis. Deep learning features between “Worse” and “Improved” outcome categories were significantly different for three radiological signs and one diagnostic (“Consolidation”, “Lung Lesion”, “Pleural effusion” and “Pneumonia”; all *P* < 0.05). Features from the first CXR of each pair could correctly predict the outcome category between “Worse” and “Improved” cases with 82.7% accuracy.

**Conclusion:** CXR deep learning features show promise for classifying the disease trajectory. Once validated in studies incorporating clinical data and with larger sample sizes, this information may be considered to inform triage decisions.

## INTRODUCTION

The current outbreak of Severe Acute Respiratory Syndrome Coronavirus 2 (SARS-CoV-2) and the subsequent pandemic of coronavirus disease (COVID-19) is imposing a substantial stress on healthcare systems worldwide. In the majority of COVID-19 cases admitted to intensive care units (ICU) for respiratory distress and hypoxaemia, endotracheal intubation and ventilation are the main treatment options. The high number of infected patients has highlighted the need for more precise decision support systems for determining the need and prognosis after ventilation, especially in healthcare networks where there is a risk of overwhelming system capacity. The recent surviving sepsis campaign recommendations do not make any specific recommendation about this triage decision making (1). Clinical prediction rules are therefore required to help caregivers during this delicate but necessary decision making process, and these rules should be based in part on the prognosis of possible outcomes (2).

While imaging is not indicated for diagnostic purposes in COVID-19, the use of chest radiography to inform prognosis was recommended by Rubin et al. in the recent consensus statement of the Fleischner society, published in this journal: “in a resource-constrained environment, imaging is indicated for medical triage of patients with suspected COVID-19 who present with moderate-severe clinical features and a high pre-test probability of disease” (3).

This recommendation rests on radiological findings for COVID-19, already reported in adults (4) (5) (6) (7). For CT imaging, they comprise (a) bilateral, subpleural, and peripheral ground-glass opacities; (b) crazy paving appearance (ground glass opacities and inter-/intra-lobular septal thickening); (c) air space consolidation; (d) bronchovascular thickening; and (e) traction bronchiectasis. COVID-19 appearance on CXR was reported more recently, with a handful of reports focusing specifically on anterior-posterior (AP CXR) at the bedside, the most common form of imaging in ICUs. CXR may be normal in early or mild disease, but commonly shows abnormal findings in patients requiring hospitalization, in 69% of patients at the time of admission, and in 80% of patients sometime during hospitalization (8). Most frequent CXR findings are consolidation (59 %) and ground glass opacities (41 %)(8) (9), with a peripheral and lower zone distribution, that are commonly bilateral or multilobar and that tend to be patchy and asymmetric. Pneumothoraces are rare. The main finding over time on CXR was consolidation (8). These findings are not specific however, being similar to other causes of coronavirus and other viral pneumonias (10). Given the critical nature of the triage decision, it is imperative that as much relevant information as possible be extracted from all available data. This information can help in assessing the risk of mortality, determine priority for initiating ventilation, determine improvements in condition and predict probable clinical trajectory. All of these must be considered in the intervention decision (11). We postulate that AP CXR images may provide such additional information, beyond simply assessing disease spread, in the form of radiomics-like features; and hypothesize that deep learning can extract these features in a reproducible and quantitative manner.

Towards this goal, we present our initial accuracy tests at tracking and predicting radiological evolution in a series of COVID-19 cases for a deep learning system adapted to extract features from AP CXR.

## MATERIALS AND METHODS

### Study design

This is a retrospective study of a large dataset of CXRs and one convenience series of COVID-19 cases, both open access. This study is conducted and reported based on the STARD criteria (12).

### Ethics

The study was approved by the ethics and research review board of our institution [Information withheld to preserve blinding].

### Dates of Study

The study was performed between March 15, 2020 and April 9, 2020.

### Training, test, and validation sets

*Training set:* We used as training set the open “CheXpert” chest X-ray dataset from Stanford Hospital, comprised of 224,316 X-ray images taken from 65,240 unique patients (aged 60.4 ± 17.8 years (mean ± standard deviation); 132,636 CXRs from men / 90,777 CXRs from women)(**Table 1**)(13). The CheXpert database was originally extracted from the Stanford Hospital PACS system with the assistance of text mining from the associated radiological reports using natural language processing. The dataset includes both posterior-anterior, anterior-posterior and lateral images. None were from COVID-19 positive patients.

**Table 1.**
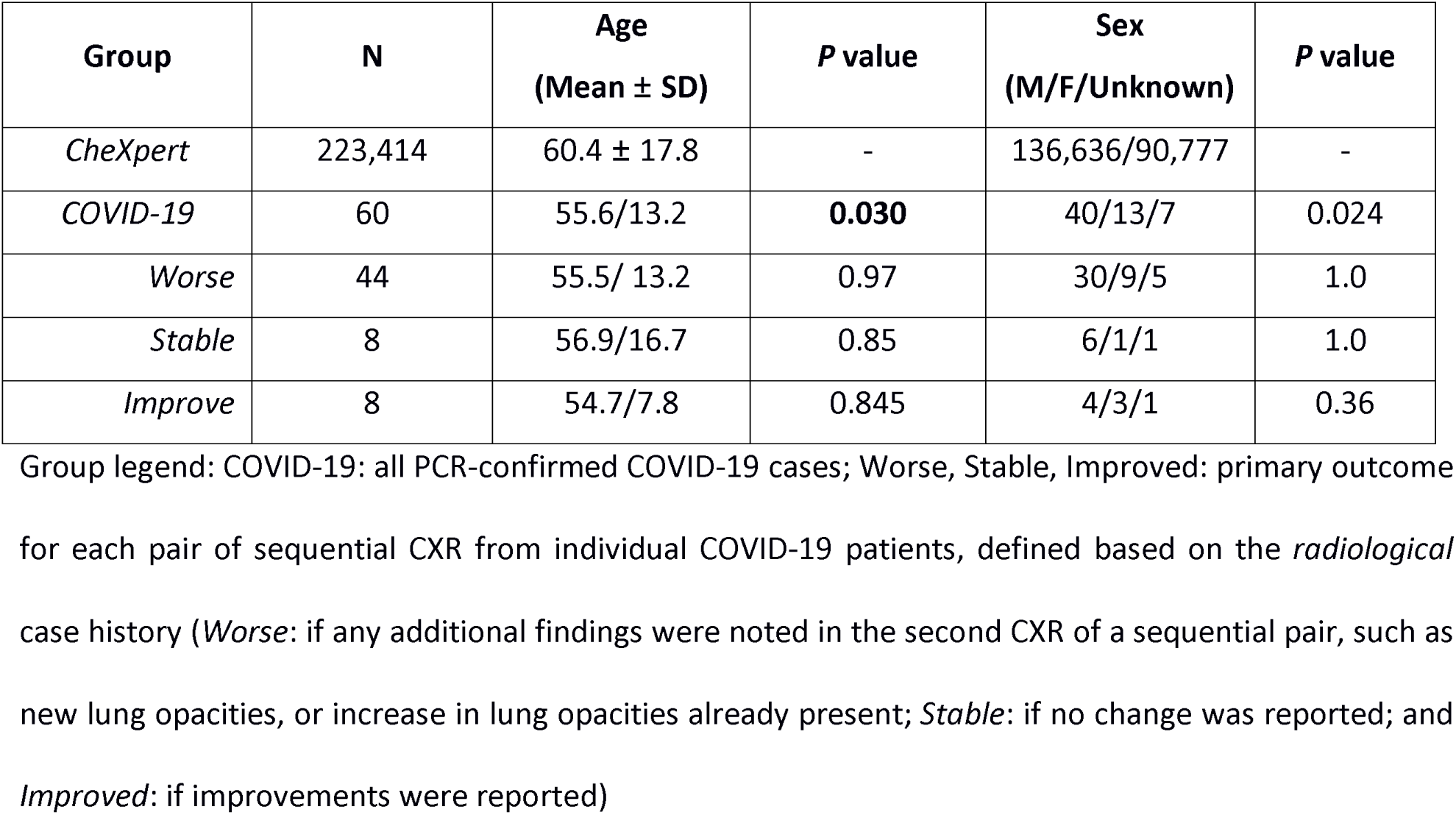
– Group demographics information

| Group | N | Age<br>(Mean ± SD) | P value | Sex<br>(M/F/Unknown) | P value |
| --- | --- | --- | --- | --- | --- |
| *CheXpert* | 223,414 | 60.4 ± 17.8 | - | 136,636/90,777 | - |
| *COVID-19* | 60 | 55.6/13.2 | **0.030** | 40/13/7 | 0.024 |
| *Worse* | 44 | 55.5/ 13.2 | 0.97 | 30/9/5 | 1.0 |
| *Stable* | 8 | 56.9/16.7 | 0.85 | 6/1/1 | 1.0 |
| *Improve* | 8 | 54.7/7.8 | 0.845 | 4/3/1 | 0.36 |

Group legend: COVID-19: all PCR-confirmed COVID-19 cases; Worse, Stable, Improved: primary outcome for each pair of sequential CXR from individual COVID-19 patients, defined based on the *radiological* case history (*Worse*: if any additional findings were noted in the second CXR of a sequential pair, such as new lung opacities, or increase in lung opacities already present; *Stable*: if no change was reported; and *Improved*: if improvements were reported)

*Validation set:* The validation set (*n =* 234) for deep learning feature extraction was selected at random within the 500 validation set studies that forms part of the CheXpert dataset (https://stanfordmlgroup.github.io/competitions/chexpert/)(13). The latter was composed of randomly sampled studies from the full dataset with no patient overlap. Three board-certified radiologists from the CheXpert team individually assigned radiological findings and diagnoses to each of the studies in this validation set.

*Test set:* The test set for COVID-19 was curated from a convenience sample of 40 cases (aged 56 ± 13) years; 23 men, 10 women, seven not reported)(**Table 1**) with sequential AP CXRs accessible in two open access repositories, the Italian Society for Medical and Interventional Radiology (https://www.sirm.org/category/senza-categoria/covid-19/) and the MILA COVID-19 image data collection (https://github.com/ieee8023/covid-chestxray-dataset/). Italian reports were translated to English by one author (F.V.). Care was taken to eliminate double entries between the datasets. A full list of cases, including links to original sources, is included in **Supplementary Material 1**.

### Inclusion/Exclusion/Eligibility

*Training and Validation set*: adult participants having visited Stanford Hospital who underwent CXR for any clinical presentation between October 2002 and July 2017 in both inpatient and outpatient settings (13).

*Test set*: adult participants initially admitted to emergency departments or ICUs for COVID-19 with sequential AP CXRs and access to summary radiological data.

### Index test and reference standard

The canonical index test for confirmation of COVID-19 was a positive polymerase chain reaction test. By patient, sequential AP CXRs were grouped into pairs. There was a total of 60 such pairs, given that some patients received more than two CXRs. The primary outcome for each pair of sequential AP CXRs was a categorical classification of radiological evolution (“Worse”; “Stable”; “Improved”) and defined based on the radiological case history provided with the open dataset as well as the images themselves **(Supplementary Material 1)**. The history was performed by certified radiologists at the centers providing cases. The categorical classification was done by two authors ([N.D.] (25 years practice); [S.L.] (fourth year residency) for indications regarding radiological outcome. If, when compared to the first CXR of a pair, any additional findings (e.g. new lung opacities, or increase in lung opacities already present) were noted in the second CXR, then the pair was categorized as “Worse”. If no change was reported, it was labeled “Stable”; and if improvements were described, the category was “Improved”. In the case of discrepancy between authors’ reading, a tie-break was provided by the lead study author. There were 44 pairs of successive CXR studies in the “Worse” outcome category; eight in the “Stable”; and eight in the “Improved” outcome categories **(Table 1)**. Mean age for the Worse outcome group was 55.5 ± 13.2 years (mean ± standard deviation)(30 men, nine women, five not reported); for the Stable group 56.9 ± 16.7 years (six men, one women, one not reported); and the Improved group 54.7 ± 7.8 years (four men, three women, one not reported).

### Deep learning

We trained a deep learning model for feature extraction, taking as input all single-view chest radiographs of the training CheXpert dataset (regardless of patient position) and providing as output the probability of nine radiological findings and one radiological diagnostic category (“Pneumonia”, used as a label by the CheXpert authors to represent images that suggested primary infection as the diagnosis). The findings were defined by certified radiologists in CheXpert. We removed the following radiological findings from the training set, given their irrelevance to the purpose of our study: “No Findings”; “Fracture”; “Support Devices”. We further removed “Pneumothorax”, given its low occurrence in COVID-19. We used a DenseNet121 architecture for all our experiments as it was determined by Irving et al. to achieve the best results on the CheXpert dataset (13). Images were fed into the network with pretrained weights on Imagenet with a size of 320 × 320 pixels. We used the Adam optimizer with default β-parameters of (β1 = 0.9, β2 = 0.999 and learning rate of 1 × 10^−4^ which was fixed for the duration of the training. Batches were sampled using a fixed batch size of 16 images. We used a weighted binary cross-entropy loss function to account for class imbalance and followed the U-zeroes policy from (13), replacing the uncertain findings with negative findings. We trained for three epochs, saving checkpoints every epoch and using the checkpoint with the lowest validation loss.

### Outcome prediction

We then proceeded in testing our hypothesis as follows **(Figure 1)**. First was whether deep learning features could track radiological evolution. We used the deep learning network as trained above to extract the findings probabilities from each CXR. We then computed the difference in findings probabilities between sequential CXRs in each pair and tested whether this difference was significant between outcome groups.

**Figure 1 -.**
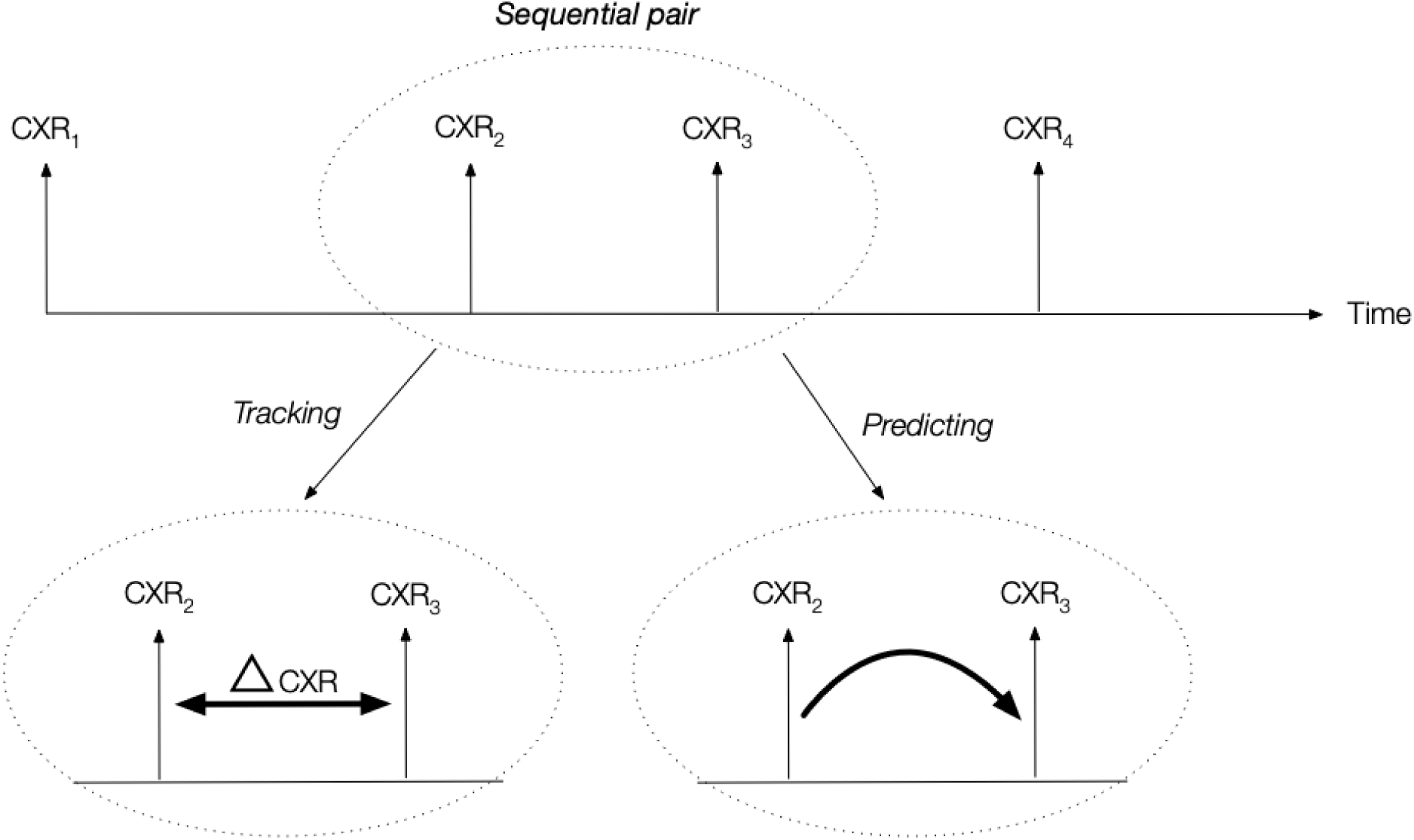
Experimental design. For each patient, the acquired CXRs formed a series of sequential pairs. For each pair (example shown for CXR_2_ and CXR_3_), an outcome was defined by judging if the radiological evolution of second CXR of the pair was worse, stable or improved compared to the first. We then tested whether the difference (ΔCXR) in radiological findings probabilities would be statistically different between outcome categories; and secondly if deep learning features from the first CXR of the pair would predict radiological evolution.

Secondly, we attempted to assess the predictive power of the extracted deep learning features, i.e. whether or not the features of the first CXR could predict the outcome category of the CXR pair (Worse or Improved)**(Figure 1)**. Instead of using the predicted findings or the findings probabilities from the deep learning network, we used the output of the last convolutional layer. We reduced the dimensionality of this feature space by selecting only the significantly different features between classes using a Chi-square test, and created a logistic regression model for the prediction of outcome category. We performed a classification using a leave-one-patient-out scheme, removing/testing all pairs associated with this patient in the learning/testing phase.

### Software

Deep learning feature extraction was done in Python using the PyTorch library (version 1.4). Our source code is available in the following GitHub repository: https://github.com/medicslab/COVID-19-public.

### Statistical Analysis

Demographics were expressed as mean (standard deviation) in years, and differences between outcome groups were tested using the SciPy library. Statistical analysis of the deep learning algorithm consisted in calculating the area under the (receiver operating) curve (AUC) for the determination of label learning on the training set for radiomics; and Mann-Whitney tests to compare results between outcome groups. We used a *P* < **0.05** threshold for significance and calculated effect size (Cohen’s *d)* for each output.

## RESULTS

### Demographics

There were no statistical differences in age and sex between COVID-19 outcome groups, however CheXpert patients were older and the proportion of males was significantly lower than the whole COVID-19 cohort (*P* < 0.05)**(Table 1)**.

### Study flowchart

There were 40 patients with at least two sequential CXRs from the open datasets **(Figure 2)**. The test was performed on 60 pairs of CXRs.

**Figure 2 -.**
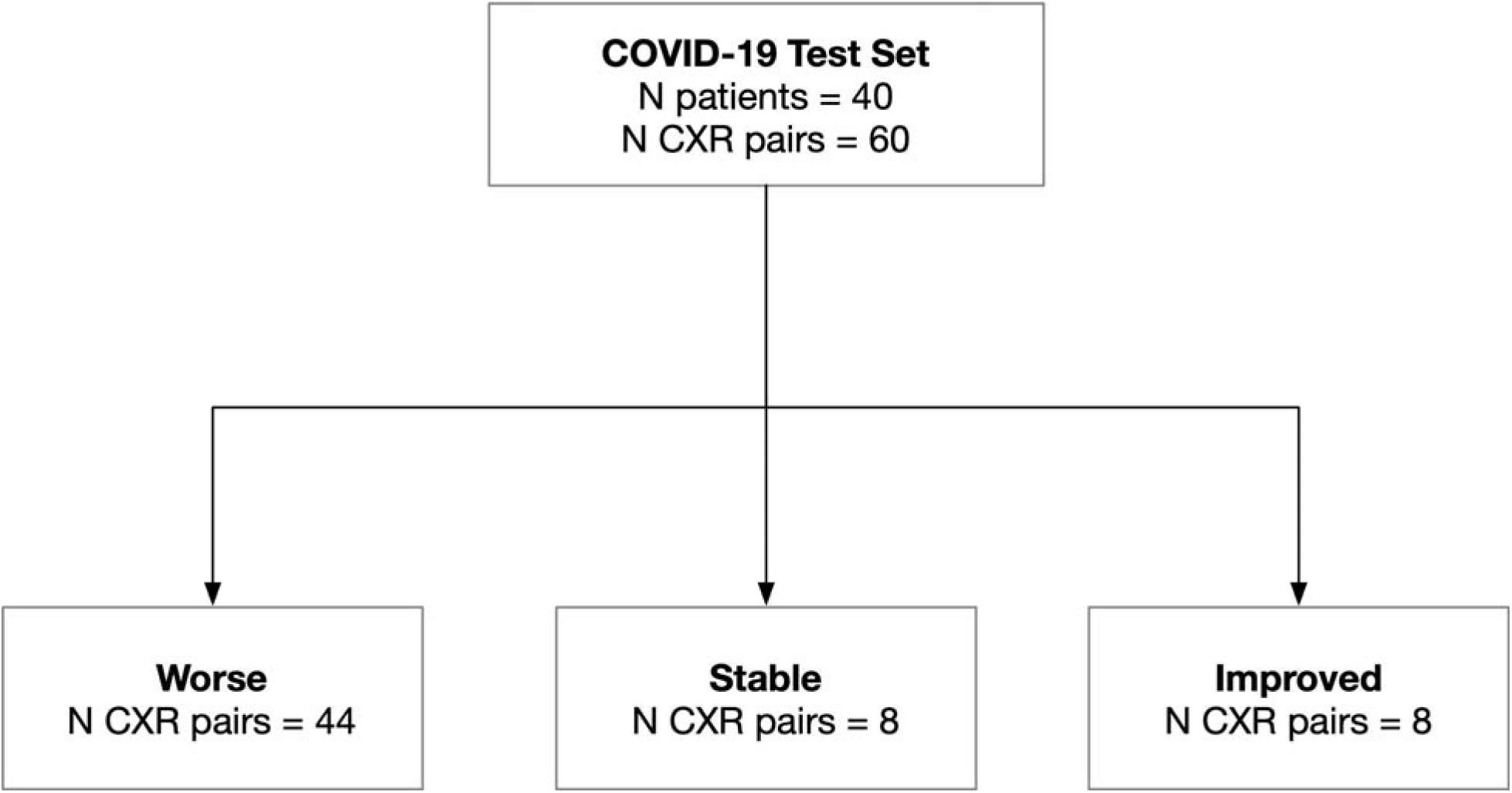
Study flowchart. Some patients may have CXR pairs in more than one outcome category.

### Deep learning feature extraction

We successfully trained the deep learning algorithm with the aforementioned architecture to extract salient radiological findings, attaining results comparable to those from the original authors of the CheXpert series **(Figure 3)** with AUCs ranging from 0.71 (“Enlarged Cardiomediastinum”) to 0.93 (“Consolidation”). We were unable to ascertain AUCs for two radiological findings (“Lung Lesion”, “Pleural - Other”) due to a lack of sufficient number of cases in the validation dataset. We generated a class-activation map for the highest-activated radiological sign (“Pneumonia:) on a random COVID-19 patient for illustrative purposes **(Figure 4)**.

**Figure 3 -.**
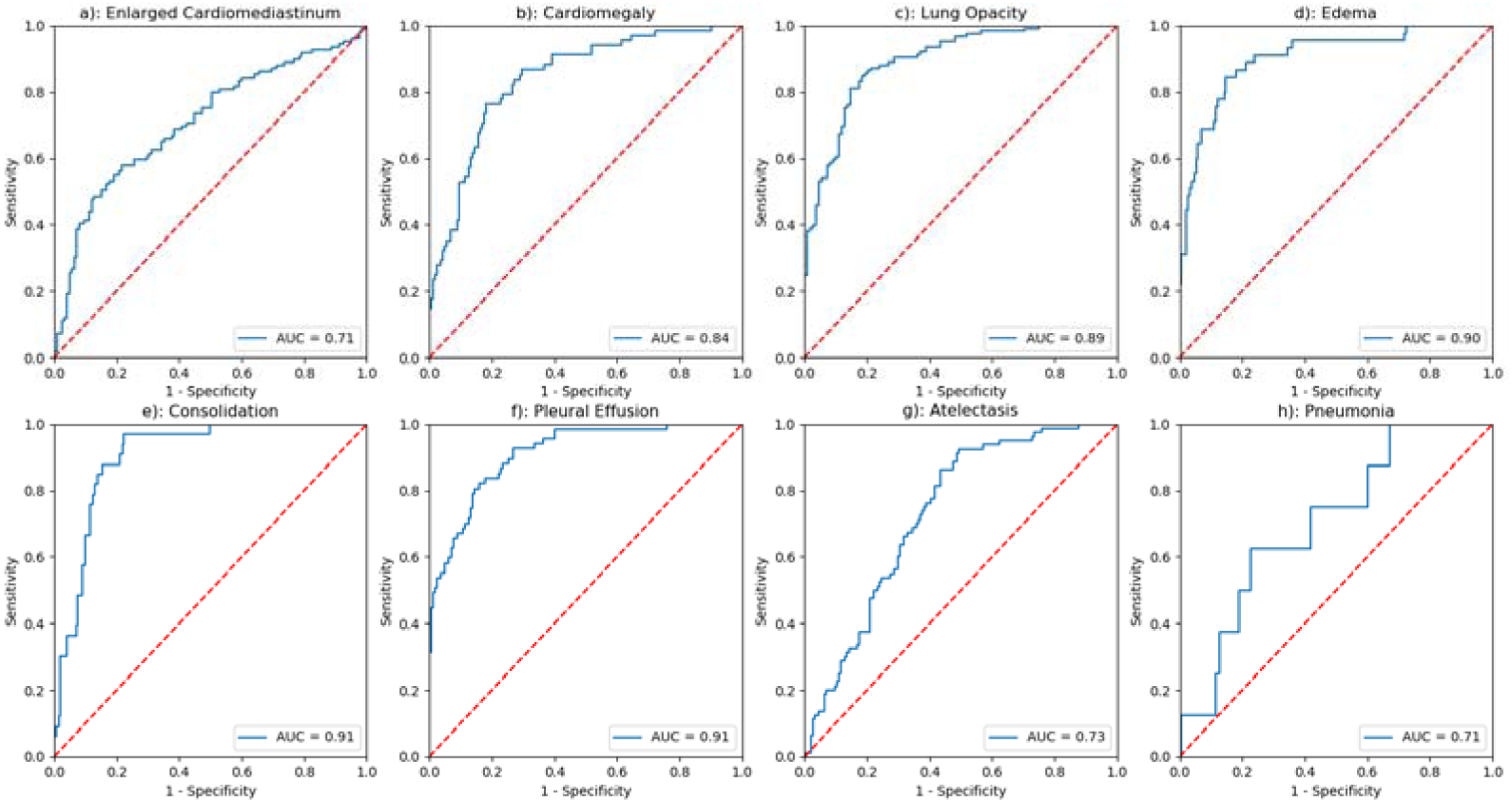
Results of the deep learning architecture trained on CheXpert for seven radiological findings (**a** to **g**) and one radiological diagnosis (**h**) on a separate 234-cases test dataset, selected at random within the 500 test set studies of the CheXpert dataset (*cf*. Irvin et al. for details). The latter was composed of randomly sampled studies from the full dataset with no patient overlap. Three board-certified radiologists individually annotated each of the studies in this test set.

**Figure 4 -.**
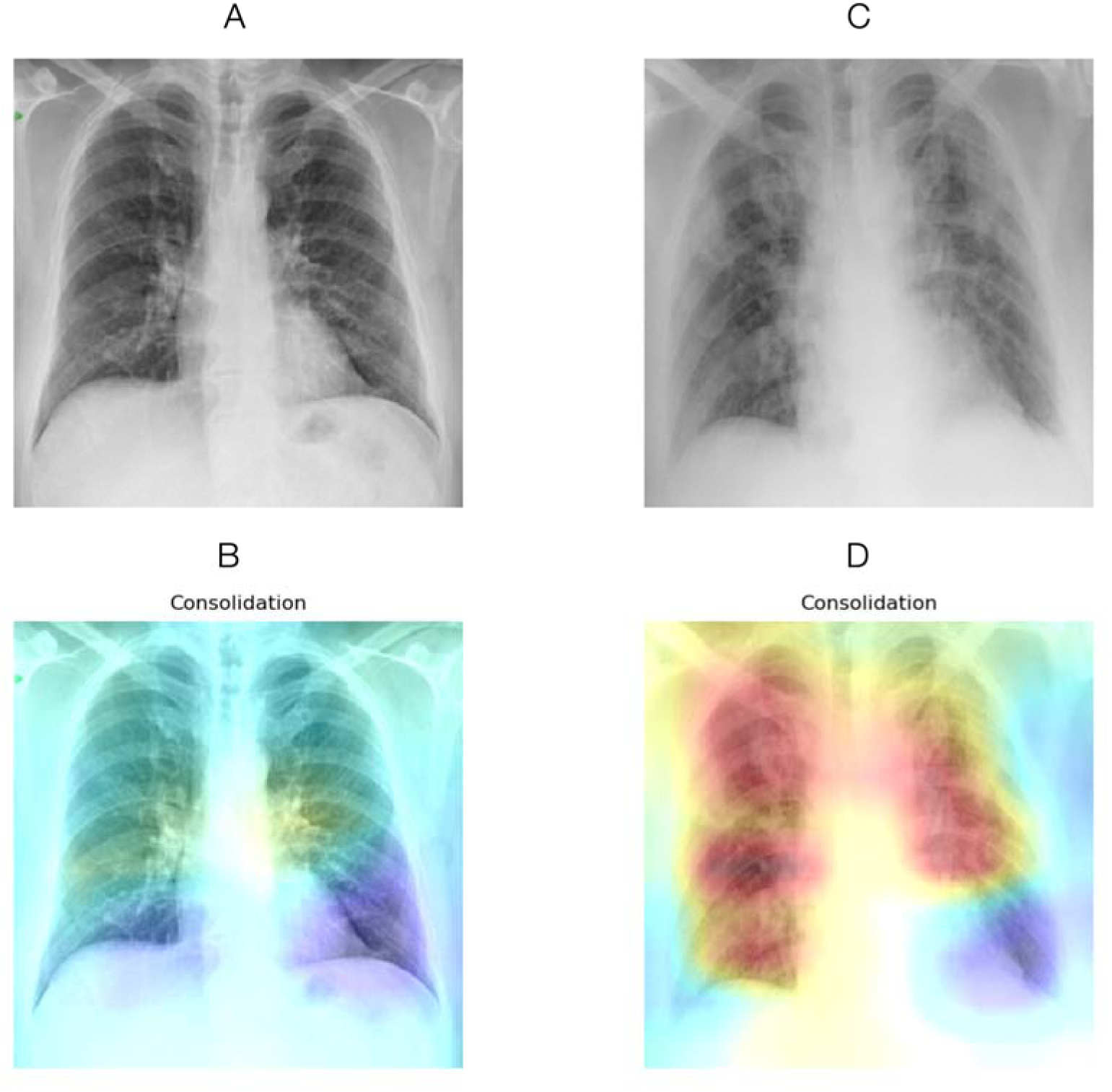
Original CXRs and class activation maps for a random patient in the COVID-19 dataset. **(A)** CXR at admission, with **(B)** overlaid activation map for the most activated radiological finding (“Consolidation”). **(C)** CXR four days later, with a worsening radiological presentation. The regions activated for the same finding **(D)** now encompass a larger area as the disease has progressed.

### Outcome prediction

Testing whether deep learning features could track disease trajectory, we applied the learned classifier to the test set, extracted radiological sign probabilities, and computed the differences between sequential CXRs. The four main findings related to COVID-19 are shown in Figure 5 for each outcome group. There were significant inter-group differences (Worse vs. Improved outcome categories; Mann-Whitney *P* < 0.05) for four radiological findings and diagnoses (“Consolidation”, “Lung Opacity”, “Pleural effusion”, and “Pneumonia”), and a significant difference only for the “Pleural effusion” sign between Worse vs. Stable groups (Mann-Whitney *P* < 0.05; **Table 2)**. The Cohen’s d effect sizes for the Worse vs Improved comparison were “Consolidation”: 0.791; “Lung Opacity”: 0.783; “Pleural effusion”: 0.479, and “Pneumonia”: 0.568. For the Worse vs. Stable case, the effect size of “Pleural effusion” was 0.764. Testing whether deep learning features could predict future outcome, the last convolutional layer was reduced from 1,024 to five features using Chi-square tests (*P* < 0.05). These features were fed to the logistic regression model. Using a leave-one-patient-out cross-validation, performance measures were: accuracy: 82.7% (confidence interval (CI): 69.7% to 91.8%); sensitivity 86.4% (CI: 72.6% to 94.8%); specificity 62.5% (CI: 24.5% to 91.5%); positive likelihood ratio: 2.3 (CI: 0.93 to 5.68); negative likelihood ratio 0.22 (CI: 0.09 to 0.55); positive predictive value: 92.7% (CI: 83.7% to 96.9%); and negative predictive value: 45.4% (CI: 25.0% to 67.6%). Reversing the order of the pairs (i.e. trying to predict the first CXR using the second of the pair) reduced accuracy to 59.6%, as expected close to chance.

**Figure 5 -.**
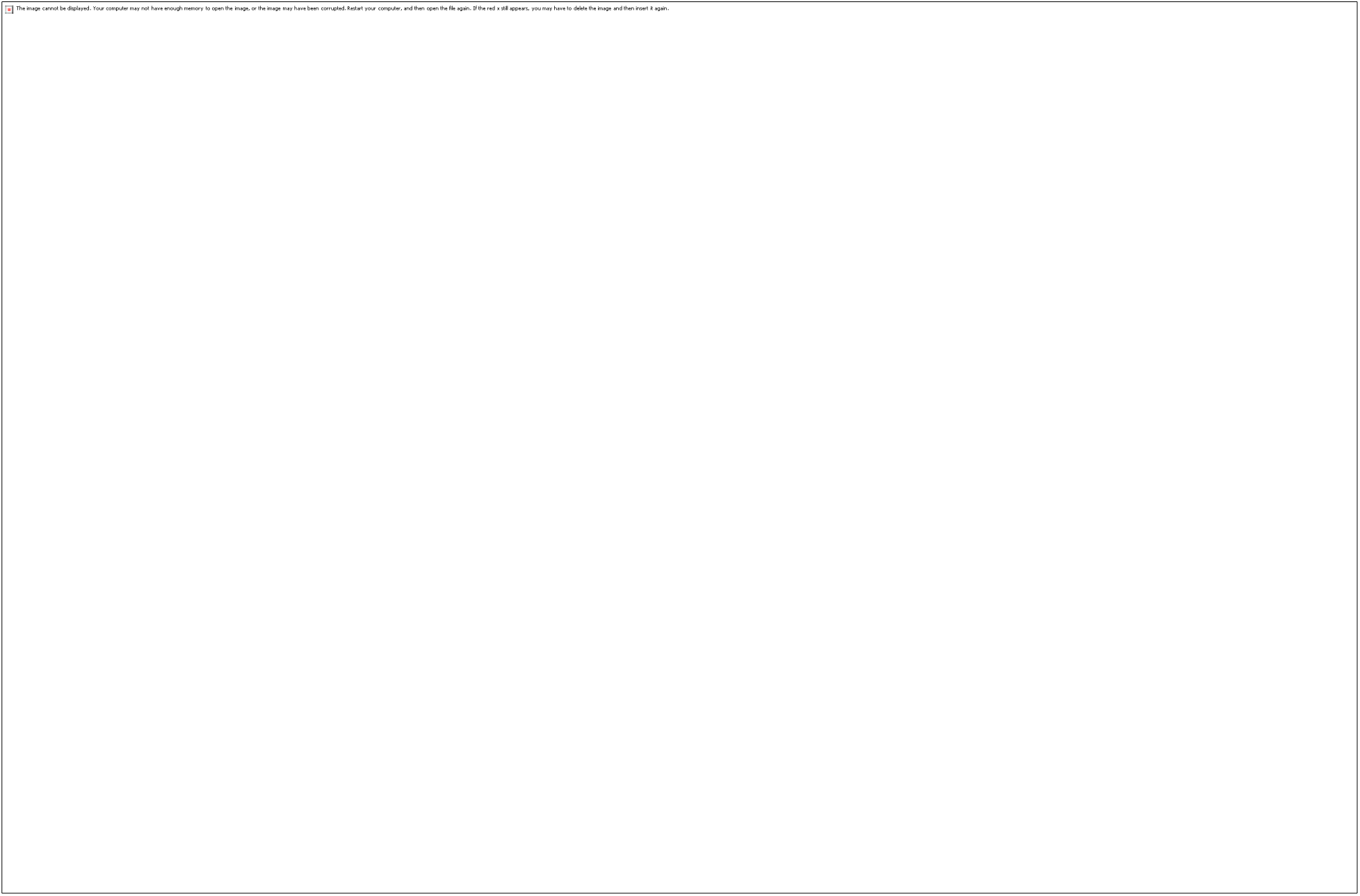
Boxplots show significant inter-group differences between Worse vs. Improving (*P* < 0.05, indicated by symbol *) in deep learning feature probabilities associated with three imaging findings: “Consolidation”, “Pleural effusion”, “Lung opacity”, and diagnosis of pneumonia.

**Table 2.** – Deep learning feature differences between outcome groups (Mann-Whitney)

|  | Worse vs. Improve | Worse vs. Stable | Stable vs. Improve |
| --- | --- | --- | --- |
| Rx sign/Diagnosis | P value<br>Cohen's d | P value<br>Cohen's d | P value<br>Cohen's d |
| *Consolidation* | **0.0171**<br>**0.791** | 0.184<br>0.533 | 0.0946<br>-0.503 |
| *Lung Opacity* | **0.0278**<br>**0.783** | 0.1908<br>0.508 | 0.186<br>-0.417 |
| *Pleural effusion* | **0.0412**<br>**0.479** | **0.0109**<br>**0.764** | 0.3183<br>0.52 |
| *Pneumonia* | **0.0232**<br>**0.568** | 0.3378<br>0.391 | 0.0946<br>-0.378 |

## DISCUSSION

### Summary

Triage decisions to decide if and when patients should be admitted in the ICU and mechanically ventilated during the current COVID-19 pandemic must be based on sound ethical guidelines and all available prognostic evidence.

We hypothesized that deep learning analysis of baseline CXR and longitudinal changes in feature probabilities could provide objective information to help in these triage decisions. To this end we needed first to prove the ability of deep learning of assessing imaging features and predicting imaging outcomes related to the disease. Consequently, we used a deep learning architecture, pre-trained on a large CXR dataset, and able to learn image features related to nine radiological signs and one pneumonia diagnosis. We applied this algorithm to a series of sequential images from patients with suspected or proven COVID-19. The algorithm was able to significantly detect changes in the images related to either a worsening or improving outcome for the patient and predict the category from the first CXR with reasonably high accuracy (>80%).

### Findings and implications for practice

We found that the proposed deep learning architecture was able to derive meaningful feature classes from a large yet disparate number of images. In effect the CheXpert dataset was not curated specifically for pneumonia; images were acquired in a variety of positions (e.g. anterio-posterior, posterior-anterior, and lateral views); and there were a number of non-pathologically related artefacts (e.g. various devices creating image shadows). Yet, it proved robust at extracting those deep learning features that best correlated to the radiological findings in the validation and test sets, the latter only composed of AP CXRs. The class activation maps of Figure 3 are indicative of the process and show that the deep learning architecture is correctly focusing on relevant areas.

The value of the deep learning features to inform triage decision making however lies not so much in the identification of radiological findings; this task is being done by radiologists themselves in the course of their duty. Rather, it centers on the ability to extract image features, distributed over the image, that may prove salient at the task of predicting outcome. These may be subtle, counter-intuitive, and therefore not part of the usual radiological diagnostic checklist or report; be subject to inter-reader variability; or couched in language that would vary between readers and centers. By quantitatively calculating these features, the model provides objective, repeatable estimates that may have better predictive ability than the binarized appraisal of disease status as exemplified in clinical scores such as the SMART-COP (“multilobar: yes/no”)(14).

### Study Limitations

This study has some limitations. First, the small size of the test dataset, which inevitably must be augmented to avoid potential bias, most notably case selection, and to confirm generalizability. However, this study represents a proof of concept whose predictive performance can be reassessed as the research community shares additional cases of COVID-19-positive CXRs. Second, the time duration between sequential CXRs was not uniform, which may have diminished the appraisal of the features’ sensitivity to change and the predictive ability of our model. Further, the design of the study is retrospective. However, as the pandemic unfolds, new clinical and radiological data will be continuously incorporated in the test set from the open source repositories, and in future cases from the authors’ institutions, which will truly test generalizability and solve most of these limitations. The authors would be grateful to any reader that would be willing to contribute to this effort.

To a degree, this report has demonstrated that deep learning features can track radiological progression in COVID-19 but also predict temporal evolution, adding evidence to the conceptualization that there is directional information in static x-rays allowing this prediction. It should be restated however that the reference standard was categorization of *imaging* rather than *clinical* outcomes, such as duration of ICU stay or mortality. Hence, it remains to be determined whether these features can track clinical, rather than radiological, progression. Further studies should therefore assess the added value of deep learning features in clinical decision making using multivariate models incorporating additional variables such as vital signs, oxygenation and ventilation parameters, and assessment of imaging data.

### Conclusion

We found that the results were sufficiently convincing to warrant further consideration of deep learning features being incorporated in a clinical prediction rule to support clinicians in making triage decisions. This being said, triage decision making and decisions to institute mechanical ventilation will not only rely on such prognostic decision rules. Shared decision making integrating the best available prognostic models, clinician experience and patient values and preferences about life-sustaining therapies will also be paramount in making these very difficult decisions. Depending on the phase of the COVID-19 viral pandemic, decisions may unfortunately only be based on prognosis, the ethical principle of social justice and availability of mechanical ventilation.

## Data Availability

All data are open access (references in manuscript)

https://stanfordmlgroup.github.io/competitions/chexpert/

https://www.sirm.org/category/senza-categoria/covid-19/

https://github.com/ieee8023/covid-chestxray-dataset/

https://github.com/medicslab/COVID-19-public

## ACKNOWLEDGMENTS

We would like to sincerely thank all patients, members of the Italian Society for Medical and Interventional Radiology and MILA groups for aggregating and/or releasing data. We would further like to thank K. Duchesne (CERVO Brain Center), as well as F. Alù, F. Miraglia, and A. Orticoni from the Brain Connectivity Laboratory of IRCCS San Raffaele Pisana, Rome, Italy for help in data collection and translation. This study has been financed by a COVID-19 Pilot Project grant from the Quebec Bio-Imaging Network as well as concurrent funding from a Discovery Award to the primary investigator (S.D.) from the National Science and Engineering Research Council of Canada. There are no relevant conflicts for the authors for this study.

## Abbreviations

AP: : anterior-posterior
AUC: : area under the (receiver operating) curve
COVID-19: : coronavirus disease
CXR: : chest X-ray
ICU: : intensive care unit

## Notes

### Competing Interest Statement

The authors have declared no competing interest.

